# Large household reduces dementia mortality

**DOI:** 10.1101/2022.01.02.22268623

**Authors:** Wenpeng You, Maciej Henneberg

**Author notes:** Corresponding Author: Wenpeng You, Address: Adelaide Medical School, The University of Adelaide, Adelaide, SA 5005 Australia.

## Abstract

**Background:** Large households/families create more positive psychological well-being which may offer a life course protection against dementia development and deliver more comprehensive healthcare to dementia patients.

**Methods:** Dementia specific mortality rates of the 183 member states of World Health Organization were calculated and matched with the respective country data on household size, Gross Domestic Product (GDP), urban and ageing. Scatter plots were produced to explore and visualize the correlations between household size and dementia mortality rates. Pearson’s and nonparametric correlations were used to evaluate the strength and direction of the associations between household size and all other variables. Partial correlation of Pearson’s moment-product approach was used to identify that household size protects against dementia regardless of the competing effects from ageing, GDP and urbanization. Multiple regression identified large household was a significant predictor of dementia mortality.

**Results:** Household size was in a negative and moderately strong correlation (r = -0.6034, p < 0.001) with dementia mortality. This relationship was confirmed in both Pearson r (r= - 0.524, p<0.001) and nonparametric (*rho*□=□-0.579, *p*□<□0.001) analyses. Regardless of the contribution of ageing, SES and urban lifestyle to dementia mortality, large household was an independent predictor of dementia mortality (r = -0.331, p <0.001) in partial correlation analysis. Stepwise multiple regression analysis selected large household as the variable having the greatest contribution to dementia mortality with R^2^ = 0.263 while ageing was placed second increasing R^2^ to 0.259. GDP and urbanization were removed as having no statistically significant influence on dementia mortality.

**Conclusions:** Independent of ageing, urbanization and GDP, large household protects against dementia mortality. As part of dementia prevention, healthcare practitioners should encourage people to increase their positive interactions with persons from their neighbourhood or other fields where large household/family size is hard to achieve.

## 1. Introduction

Dementia is an umbrella neurological syndrome resulting from more than 100 brain disorders, most common of which include Alzheimer’s disease, vascular diseases, Lewy bodies, frontotemporal disease etc. [1, 2]. Worldwide, it is estimated that 5-8% general population aged 60 and over have been diagnosed with dementia in past years [2]. Therefore, it has become one of the most common causes of dependency and disability among the old [2, 3].

Dementia produces an increasing societal burden resulting in a total economic costs of US 818 billion (1.1% of the world’s gross domestic product) in 2016 [2], which has been considered as the major challenging health issue in 21st century [4, 5]. Many health professionals do not follow the regulations or facility policies about human rights and freedom, and still consider physical or chemical restraints as the inevitable approach to control patients’ behavioural symptoms and prevent the disruption of life-sustaining therapies [2]. Furthermore, dementia patients and their families are subject to prejudice, and their life quality is affected [3].

Since Alzheimer’s disease has been associated with dementia in 1906, enormous investment has been allocated for studying dementia prevention and treatment [6, 7]. Ageing and family history are associated with genetic background which are easily recognized as the irreversible risk factors for dementia [2, 8, 9]. The environmental and behavioural risk factors, such as sedentary lifestyle, lower socioeconomic status (SES) and unhealthy diet have been circumstantially postulated as the risk factors for dementia [8, 10-13]. Studies have consistently revealed that negative psychological functioning, such as depressive symptoms [14] and neuroticism [15] are the risk factors for dementia. However, to date, the aetiology and pathology of dementia are still not well understood and there is still no treatment currently available to cure dementia or to reverse its progressive course [2].

Human species had lived in small hunter-gatherer groups for millions of years before they started to live in large scale societies some ten thousand years ago [16]. During the hunting and gathering period, humans have well adapted early to cooperative breeding [17, 18], and then evolved alloparental care [19]. Therefore, human’s millions of years of adaptation suggest that biological foundations of human love have genetically shaped humans for flourishing in in small communities [20, 21]. However, in the past hundreds of years, human sorceries were industrialized quickly. The rapid industrialization has made most people grow up in core families with few siblings, which is different from how humans had adapted for flourishing. Such discrepancies or mismatches have been associated with mental health in human population [22, 23].

It is well researched that positive psychological well-being has been implicated in health across adulthood [24]. Household creates a social environment which is salient to maintain health for the co-residential members. On a daily basis, the individual members encounter this environment, play their social role and enjoy the social relations [25]. Moreover, studies also showed that large household offers the residents the subjective happiness [26] leading to low risk for residents to develop various cancers [27]. Studies also suggested that subjective happiness was associated with mental health significantly stronger than with physical health in disability people [28] and hospital patients [29]. A recent study revealed that greater household size has the protecting role against children developing mental health disorders [30]. This has directed us to identify possible contributing factors for dementia from the evolutionary perspective. Therefore, in this study, we assessed, from a global perspective, whether large household has the inhibitory role in lowering the risk for the residents to develop dementia using empirical population level data obtained from international organizations.

## 2. Materials and Methods

### 2.1 Data Sources

The population level data were collected for this ecological study.

1. Population specific dementia mortality rate (DMR, per 100,000) was calculated as the dependent variable. The comprehensive and comparable assessment of country specific number of deaths due to “Alzheimer disease and other dementias” and the country specific total population were provided by the WHO Global Health Estimates [31]. Details regarding data, methods and cause categories are described in the WHO Technical Paper [32]. The formula for dementia mortality calculation is below:

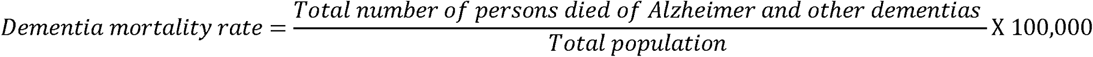
2. The population specific household size was extracted from the United Nations Booklet as the predicting/independent variable [33]. The household is a fundamental socio-economic unit in human societies. It consists of one individual or a group of people, regardless of whether there are any kinship ties, living together for sharing food, shelter and other daily life essentials. Therefore, household refers to people living together in a housing unit who may or may not be family members. Household relations are usually characterized with family relationships because they are invested with the powerful norms, histories, and emotions which originated from family [25, 34-36]. Therefore, in this study family and household are used interchangeably. Considering the majority of dementia patients are cared for at home which is called “informal” care, household size may represent the level of care which the patients can receive.
3. The WHO published life expectancy at 60 (Life e(60)) was selected as a potential confounder [37]. Ageing has been a well-known significant risk predictor of dementia. Most of the studies include 65 years age as the start of ageing for reporting the prevalence and incidence of dementia. However, the United Nations generally use 60+ years to refer to the older population, and it takes years for dementia associated symptoms and signs to appear because development of most types of dementia is slow and progressive [38, 39]. Therefore, in this study, Life expectancy at age 60 years (e60) was considered as the indicator of ageing.
4. The World Bank published data on Gross Domestic Product (GDP) and urbanization were also included as the potential confounder [40]. GDP was expressed in per capita purchasing power parity (PPP in current international $) in 2010. SES has been associated with prevalence and mortality rates of dementia, and also with regional variation of dementia prevalence. GDP PPP was included as the confounding factor because it relates to the levels of healthcare service which affects the mortality rate. Urbanization was measured with the percentage of total population living in urban communities in 2010. Urbanization represents the demographic trend in which more and more population has become concentrated in urban communities. It entails air pollution, consumption of food with few nutritional benefits, but energy dense, high levels of salt, fat, sugar and alcohol. Urbanization is also associated with less physical exercise, obesity and overweight. Therefore, urban living has been considered as a complex risk factor for chronic diseases [41].

### 2.2 Data Selection

In order to capture as many populations as we could to increase the sample size for this study, we kept the full list of 183 WHO member states (populations) which have the data available for dementia mortality rate calculations. Population specific household size, family size, urbanization life e(65) and GDP PPP were matched for those states with dementia mortality rate data. We obtained most recent population specific dementia mortality rates (N=183) through calculation, household size (N=170), GDP PPP (N=178), Urbanization (N=183) and ageing (N=183) through extraction. Each population was considered as an individual research subject in the analysis. Therefore, numbers of populations included in analyses of relationships between variables may differ somewhat because all information was not uniformly available for all states.

All the aforementioned data were freely available from the websites of the UN agencies.

### 2.3 Data analysis

Scatter plots were produced in Excel (Microsoft^®^ 2016) to explore and visualize the correlations between household size and dementia mortality rates. Scatter plots also allowed us to assess data quality and distributions of the two variables.

Prior to correlation/regression analyses all data were log-transformed (ln) in order to reduce non-homoscedasticity of their distributions and possible curvilinearity of regressions. To assess the relationships between household size and dementia mortality rate in different data analysis models, the analysis proceeded in four (4) steps.

1. Pearson’s and nonparametric correlations (Spearman’s rho) were used to evaluate the strength and direction of the associations between household size and all other variables, including independent variables and competing variables.
2. Partial correlation of Pearson’s moment-product approach was used to assess the relationship between household size and dementia mortality rate while we controlled for ageing, GDP PPP and urbanization which have been commonly considered as the contributing factors of dementia. We alternated the four variables (DMR, ageing, GDP PPP and urbanization) as the independent predictor to explore its relationship to DMR while keeping all the other three variables statistically constant. This allowed us to analyse and compare the levels of correlations between DMR and four potential risk factor while controlling for the other three variables [42, 43]. Subsequently, we alternately controlled for each variable as the potential confounder for analysing if and how much it could explain the correlation between DMR and each of the three variables. Fisher’s r-to-z transformation was performed to test significance of differences between correlation coefficients.
3. Standard multiple linear regression (enter and stepwise) was performed to visualize the correlation between DMR and each predicting factor and identify the most significant predictor(s) of DMR respectively. In order to explore if household size can partially explain why ageing, GDP PPP and urbanization were correlated with DMR, the multiple linear regression analyses were performed to determine the correlations between DMR incidence and the risk factors in two models, i.e. with and without incorporating household size as one of the predicting variables.
4. In order to demonstrate that correlation universally exists between household size and DMR regardless of these factors, populations were grouped for correlation analyses. The exploration into different correlations between household size and DMR also allowed us to compare the different levels of correlations in different country groupings. The criteria for grouping countries used the World Bank income classifications [44], WHO regions [45], countries sharing specific characteristics like geography, culture, development role or socio-economic status, like Asia Cooperation Dialogue (ACD) [46], Asia-Pacific Economic Cooperation (APEC) [47], the Arab World [47], Latin America and the Caribbean (LAC) [48], Southern African Development Community (SADC) [49] and Organization for Economic Co-operation and Development (OECD) [47]. All the population listings are sourced from their official websites for matching with the list of populations with DMR.

Pearson’s, non-parametric Spearman’s rho correlations, partial correlation and multiple linear regression (enter and stepwise) were computed with SPSS v. 27 (SPSS Inc., Chicago Il USA). The significance was reported when p-value was <0.05, but the significance levels of p < 0.01 and p<0.001 were also reported. Regression analysis criteria were set at probability of F to enter ≤ 0.05 and probability of F to remove ≥ 0.10. The raw data were used for scatter plots in Excel^®^ 2016.

## 3. Results

Figure 1 shows that the relationship between household size and DMR is logarithmic with a negative and moderately strong correlation (r = -0.6034, p < 0.001). The non-linear relationship between household size and DMR variables identified in the scatterplots was confirmed by the subsequent analyses of log-transformed data and in nonparametric analyses.

**Figure 1:**
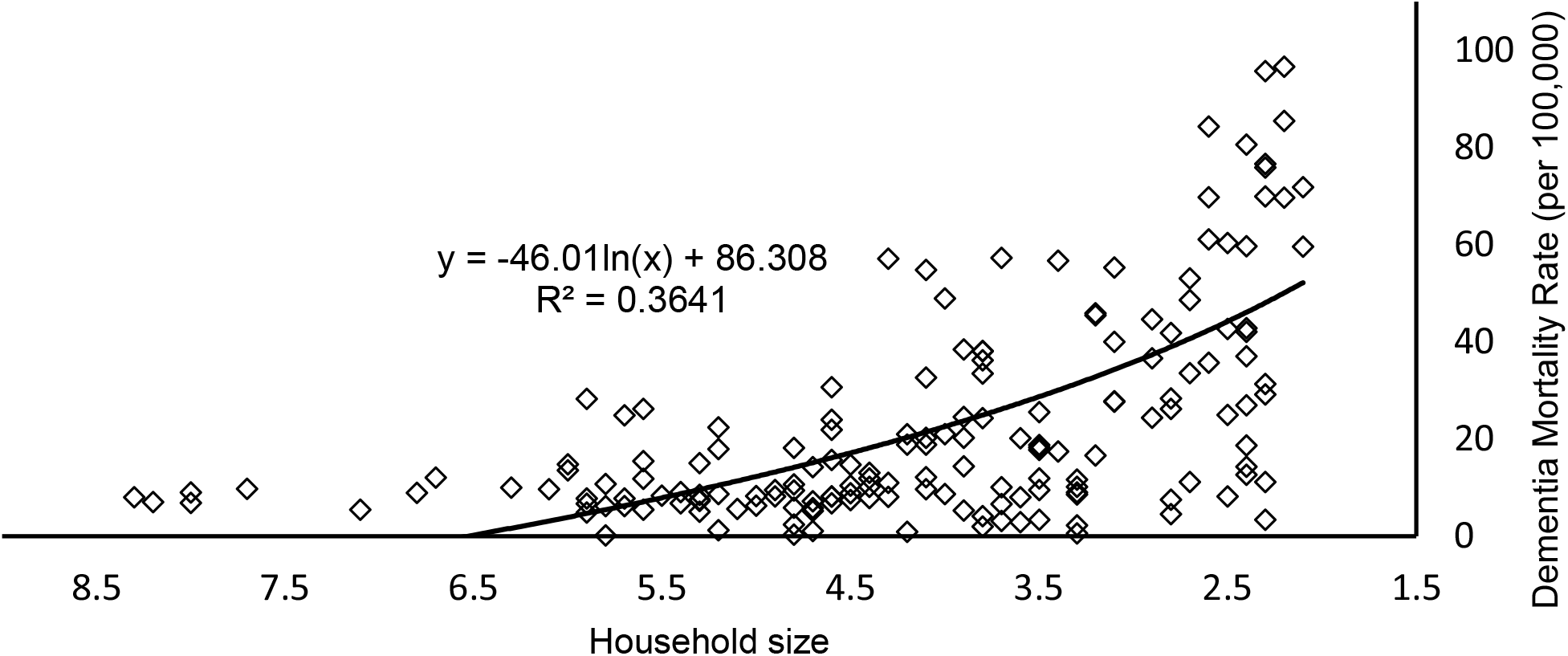
The relationship between household size and dementia mortality rate. Data source & definition: Household size (the United Nations): the number of persons who make common provision of food, shelter and other essentials for living. Dementia Mortality Rate (World Health Organization): Calculated from Global Health Estimates. Both data were not log-transformed.

Table 1 presents the relationships between all the variables (dependent and independent) in Pearson r (above the diagonal) and nonparametric (below the diagonal) analyses. Worldwide (n=169), Spearman’s rank correlation showed that household size was in significant negative correlation to DMR (*r*□=□-0.579, *p*□<□0.001). This strength and direction of relationship were similar and observed in Pearson’s *r* household size and SMR variables (r= - 0.524, p<0.001). Worldwide, non-parametric analysis showed that DMR was associated with ageing (r = 0.533, p<0.001), GDP PPP (r = 0.497, p<0.001) and urbanization (r = 0.436, p<0.001). These strengths and directions of the relationship were observed in the Pearson analysis as well (Table 1).

**Table 1.**
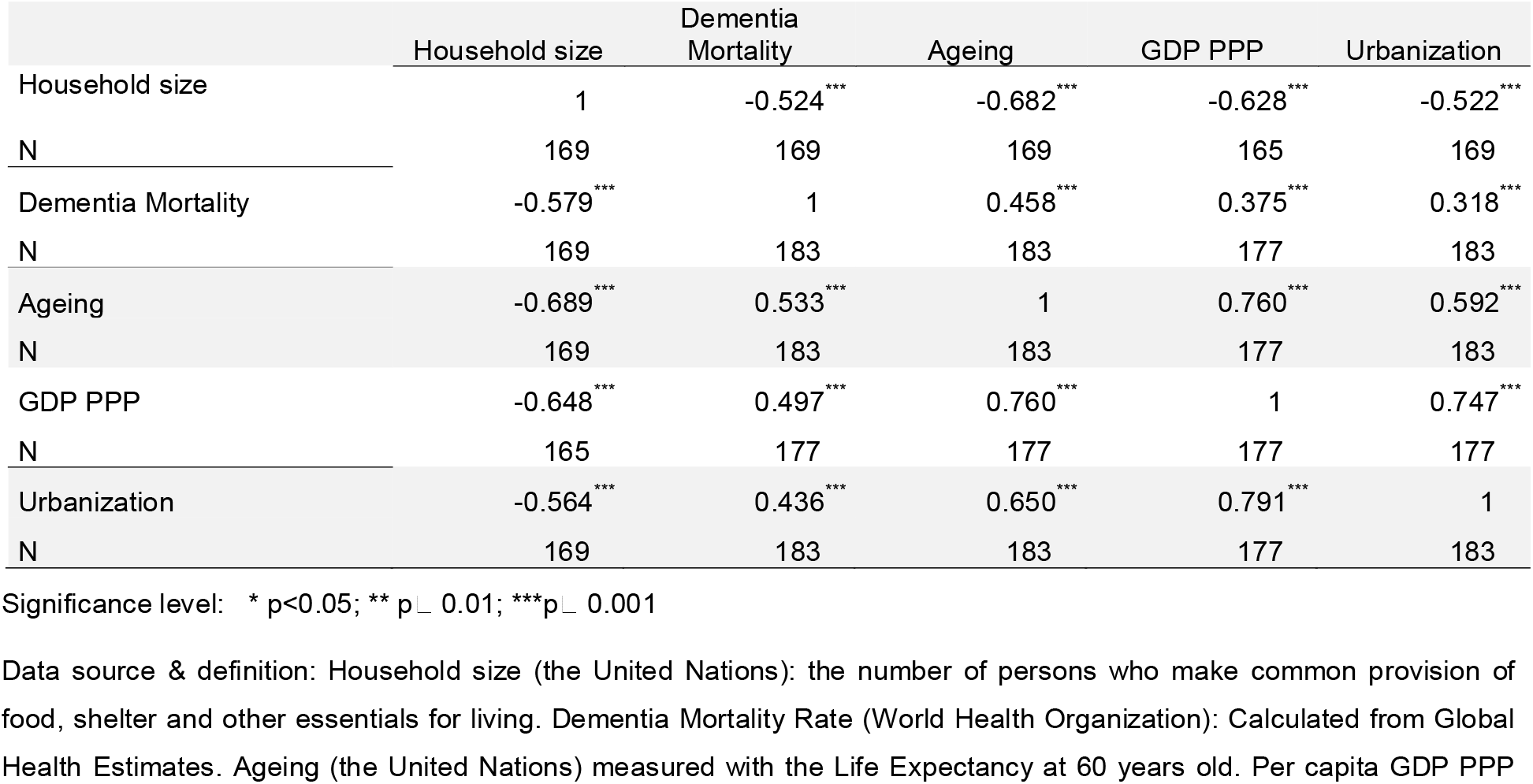
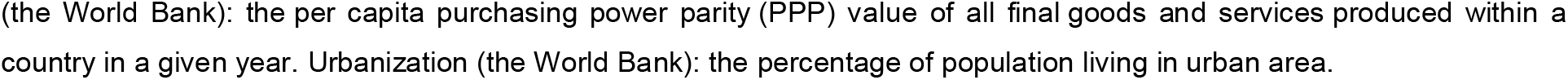
Pearson (above diagonal) & Non-parametric (below diagonal) correlation matrix for all variables.

Table 2 shows that the relationship between dependent variable (DMR) and each independent variable (household size, ageing, GDP PPP and urbanization) was examined by controlling for the other three variables in a partial correlation analysis. Household size and ageing were the only independent variables to have significant correlations (r = -0.331, p < 0.001 and 0.173, p<0.05) with DMR independent of the other three variables (Table 2). Neither the other two predicting variables (GDP or urbanization) showed a correlation with DMR incidence independent of the other three variables despite the fact that each of them (GDP and urbanization) had a significant correlation to DMR in Pearson r and non-parametric correlation analyses respectively. This suggested that household size was the independent risk factor for DMR. This determining role was proved in the subsequent Stepwise linear regression analyses (Table 3).

**Table 2.** Comparison of partial correlation coefficients between dementia mortality rate and each variable when the other three variables are controlled for.

| Household size, aging, GDP PPP and urbanization were alternated as the predicting variable for calculating its relationship with dementia mortality rate while the other three independent variables were kept statistically constant. |  |  |  |  |  |  |  |  |  |  |  |  |
| --- | --- | --- | --- | --- | --- | --- | --- | --- | --- | --- | --- | --- |
| Variables | Dementia Mortality |  |  | Dementia Mortality |  |  | Dementia Mortality |  |  | Dementia Mortality |  |  |
|  | R | P | df | r | p | df | R | p | df | r | p | df |
| Household size | -0.322 | <0.001 | 159 | - | - | - | - | - | - | - | - | - |
| Ageing | - | - | - | 0.154 | 0.051 | 159 | - | - | - | - | - | - |
| GDP PPP | - | - | - | - | - | - | -0.020 | 0.798 | 159 | - | - | - |
| Urbanization | - | - | - | - | - | - | - | - | - | 0.018 | 0.821 | 159 |
| Household size, aging, GDP PPP and urbanization were alternated as the potential confounder for exploring the partial correlations between dementia mortality rate and the other three independent variables. |  |  |  |  |  |  |  |  |  |  |  |  |
| Variables | Dementia Mortality |  |  | Dementia Mortality |  |  | Dementia Mortality |  |  | Dementia Mortality |  |  |
|  | R | P | df | r | p | df | r | p | df | r | p | df |
| Household size | -0.322 | <0.001 | 159 | - | - | - | - | - | - | - | - | - |
| Ageing | - | - | - | 0.154 | 0.051 | 159 | - | - | - | - | - | - |
| GDP PPP | - | - | - | - | - | - | -0.020 | 0.798 | 159 | - | - | - |
| Urbanization | - | - | - | - | - | - | - | - | - | 0.018 | 0.821 | 159 |

Table 3 showed if and how much household size explained the correlations of the other three variables to DMR respectively. In the enter model, when household size was not considered as one of the independent variables, ageing was the only significant predictor of DMR (β = 0.357, p < 0.001). However, when household size was incorporated as an independent variable, it ranked as the strongest predictor of DMR (β = - 0.362, p < 0.001). Ageing was still a significant predictor of DMR (β = 0.226, p < 0.05), but the correlation strength has significantly decreased (z=1.67, p<0.05). Both GDP and urbanization showed bare correlations to DMR regardless of household size inclusion.

**Table 3.** Multiple linear regression showing predicting effects of independent variables and identify the significant predictors of dementia mortality.

| **Enter** |  |  |  |  |
| --- | --- | --- | --- | --- |
| Dementia Mortality |  |  |  |  |
| Household size excluded |  |  | Household size included |  |
| Variable | Beta | Sig. | Beta | Sig. |
| Household Size | - | - | -0.357 | 0.000 |
| Ageing | 0.400 | 0.000 | 0.236 | 0.036 |
| GDP PPP | 0.043 | 0.735 | 0.063 | 0.624 |
| Urbanization | 0.039 | 0.704 | -0.076 | 0.474 |
| **Stepwise** |  |  |  |  |
| Dementia Mortality |  |  |  |  |
| Household size excluded |  |  | Household size included |  |
| Model | Variable | Adjusted R<sup>2</sup> | Variable | Adjusted R<sup>2</sup> |
| 1 | Ageing | 0.203 | Household Size | 0.263 |
| 2 | - | - | Ageing | 0.289 |
| 3 | - | - | - | - |
| 4 | - | - | - | - |
| 5 | - | - | - | - |

Significance level: \* $p < 0.05$ ; \*\* $p \leq 0.01$ ; \*\*\* $p \leq 0.001$
Data source & definition: Household size (the United Nations): the number of persons who make common provision of food, shelter and other essentials for living. Dementia Mortality Rate (World Health Organization): Calculated from Global Health Estimates. Ageing (the United Nations) measured with the Life Expectancy at 60 years old. Per capita GDP PPP (the World Bank): the per capita purchasing power parity (PPP) value of all final goods and services produced within a country in a given year. Urbanization (the World Bank): the percentage of population living in urban area.

Table 3 also shows that when household size was not included as one of the independent variables, ageing (R^2^ = 0.203) was the only variable included as the significant predictor of DMR. However, when household size was included as an independent variable, household was selected as the variable having the greatest influence on DMR with R^2^ = 0.263, while ageing was placed second increasing R^2^ to 0.289. The other variables (GDP PPP and urbanization) were removed by the analysis as having no statistically significant influence on DMR.

Table 4 presents that, in general, household size is negatively associated with DMR in different country groupings. However, the highlight of these relationships was that household size was constantly in negative correlation to DMR in economically developed country groupings, such as in the World Bank High income economics, WHO European Region and OECD.

**Table 4.**
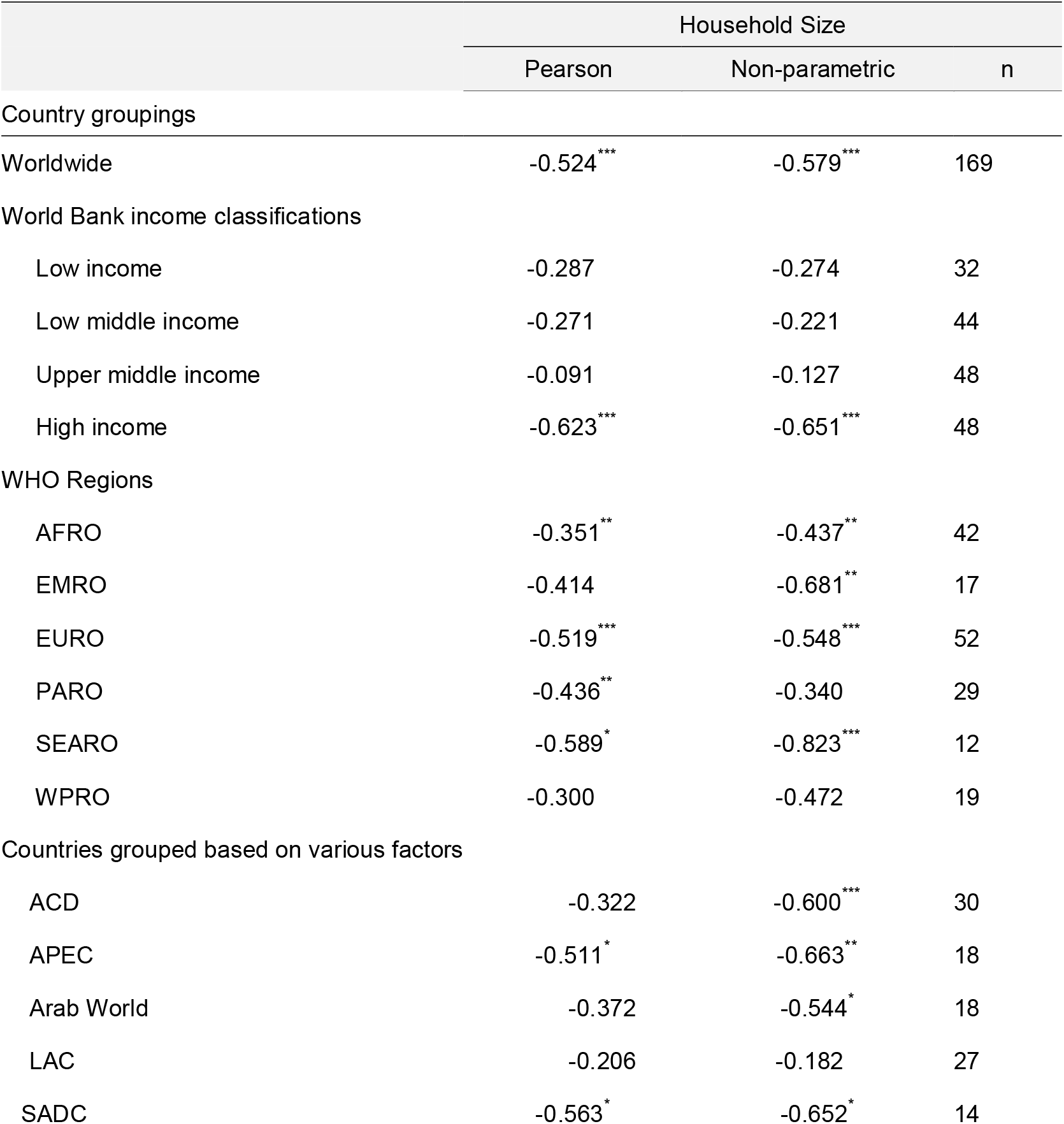
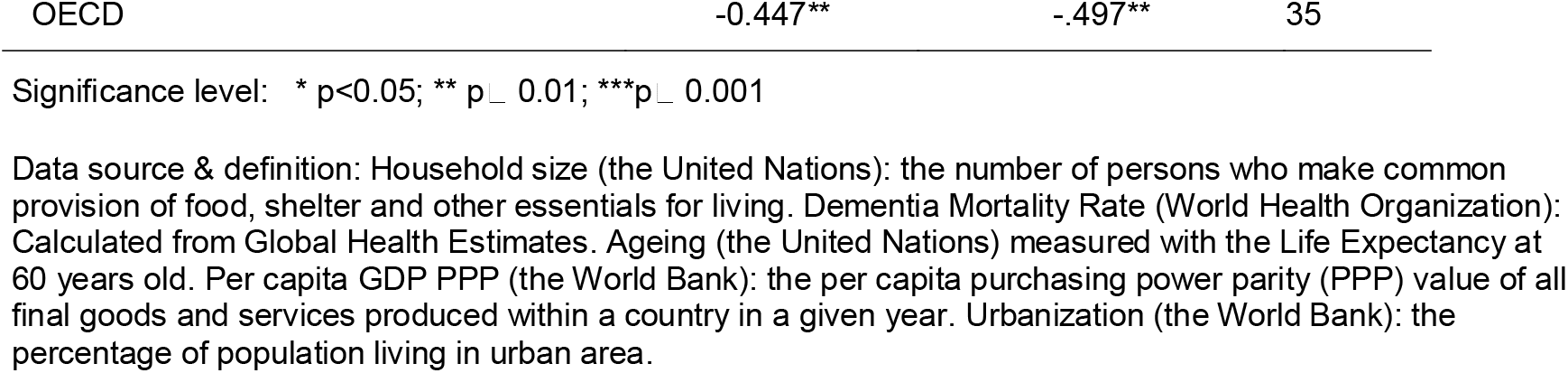
Correlation of dementia mortality to household size in different country groupings.

|  | Household Size |  | n |
| --- | --- | --- | --- |
|  | Pearson | Non-parametric |  |
| Country groupings |  |  |  |
| Worldwide | -0.524*** | -0.579*** | 169 |
| World Bank income classifications |  |  |  |
| Low income | -0.287 | -0.274 | 32 |
| Low middle income | -0.271 | -0.221 | 44 |
| Upper middle income | -0.091 | -0.127 | 48 |
| High income | -0.623*** | -0.651*** | 48 |
| WHO Regions |  |  |  |
| AFRO | -0.351** | -0.437** | 42 |
| EMRO | -0.414 | -0.681** | 17 |
| EURO | -0.519*** | -0.548*** | 52 |
| PARO | -0.436** | -0.340 | 29 |
| SEARO | -0.589* | -0.823*** | 12 |
| WPRO | -0.300 | -0.472 | 19 |
| Countries grouped based on various factors |  |  |  |
| ACD | -0.322 | -0.600*** | 30 |
| APEC | -0.511* | -0.663** | 18 |
| Arab World | -0.372 | -0.544* | 18 |
| LAC | -0.206 | -0.182 | 27 |
| SADC | -0.563* | -0.652* | 14 |

Significance level: \* $p < 0.05$ ; \*\* $p \leq 0.01$ ; \*\*\* $p \leq 0.001$
Data source & definition: Household size (the United Nations): the number of persons who make common provision of food, shelter and other essentials for living. Dementia Mortality Rate (World Health Organization): Calculated from Global Health Estimates. Ageing (the United Nations) measured with the Life Expectancy at 60 years old. Per capita GDP PPP (the World Bank): the per capita purchasing power parity (PPP) value of all final goods and services produced within a country in a given year. Urbanization (the World Bank): the percentage of population living in urban area.

## 4. Discussion

The worldwide trend of increased DMR may have multiple aetiologies, which may act through multiple mechanisms. This study not only suggested that household size may be a major factor for dementia mortality at the population level, but also showed that household size was a determining risk factor overriding risk factors, such as ageing, SES and urbanization. This study also revealed that the predicting effect of household size on dementia mortality was independent of the effects of other common risk factors, such as ageing, socio-economic status and urbanization.

Household relations are usually invested with the powerful norms, histories, and emotions that characterize family relationships [25]. From a life course perspective, they are related to the dementia risks in the essential pathways which are grounded on biological, psychological and social contributions [50]. With living in the industrialized societies, especially in the developed countries, most people grew up in core families with very limited number of playmates and little interaction with neighbours. This is different from how humans had adapted for flourishing through early cooperative breeding [17, 18], and then evolved alloparental care [19], both of which laid the biological foundations of human love which may be heritable generation by generation [20]. The mismatch between the way we live and how our ancestors did has been postulated as the risk factor for mental disorder in young generation [22, 23].

Due to lack of cure for dementia, and accurate diagnoses of specific causal factors, this has made it difficult for targeting the preventative interventions [51]. It is well established that appropriate psychological, social support and physical care have been the key strategies for the healthcare for dementia patients [52]. Studies have revealed that, psychologically, family life increases levels of purpose in life [53, 54], which may reduce by 30% risk of dementia [55, 56]. Large household may offer the residents more social engagement in life which is protective against dementia [57, 58].

Compiled 2,200 years ago, *Huangdi Neijing* has been the fundamental doctrinal source of Asian medicine. It illustrates how emotions are associated with the visceral organs which are in charge of five qi’s (translation: gas; meaning: emotions): happiness, anger, sadness, worry and fear. Among these five qi (emotions), only happiness makes the gas smooth [64], which keeps people healthy. In Western medicine, hundreds of years’ exploration of the manifestation of emotions through physiological responses (mind - body interaction) [59-61] has suggested that the formation of mental experiences (emotions) is closely associated with bodily responses [62, 63].

### 4.1 The therapeutic effects of oxytocin in dementia prevention and treatment

Oxytocin is a hormone and a neurotransmitter that is associated with social bonding, such as empathy, trust, sexual activity, group bonding and relationship-building [65]. A stream of studies in the last decade reported that oxytocin release is not only associated with giving birth [66] and lactation [67], but also with daily interactions between non-kin household residents and/or family members, such as spouses [68-70], mother and children [71], father and children [72] and co-residential household members [73-75]. Oxytocin can keep family members and household residents happy and loyal to each other [76, 77], which may bring more positive psychological well-being to the family members. Regardless of cultural backgrounds [26], people from large household, especially from the same family have more life satisfaction [26, 78] which may lead to more oxytocin production within the hypothalamo-pituitary magnocellular systems. A self-reinforcing cycle is formed between more household interactions and more oxytocin production [65].

A systematic review conducted by the researchers from the University of California concluded that the beneficial effects of oxytocin on neurological disorders may be general rather than specific through mediating their behaviours [79]. In the past decades, oxytocin has been considered a neurohypophyseal hormone which acts in social bonding, mediating stress reproduction, brain neuromodulation and central regulation of cardiovascular functioning. It has been considered a candidate for developing a new therapeutic intervention in aging and in related neuropsychiatric disorders, in particular dementia [80, 81]. Alzheimer’s disease is the most common cause of dementia. The most prevalent hypothesis about the pathology is that it occurs when the beta-amyloid (toxic protein) begins to clump around neurons in the brain. The neurons degenerate, which leads to a decrease in synaptic plasticity and ultimately to cognitive decline. Oxytocin has been considered chemical responsible for targeting the removal or reduction of beta-amyloid or improve the cognitive ability of patients. An animal model study showed that toxic beta-amyloid damaged synaptic plasticity in mice’s brains, but this beta-amyloid-induced impairment of hippocampal synaptic plasticity in mice was reversed by oxytocin in mice [82]. Interestingly, this study also revealed that oxytocin did not improve brain’s synaptic plasticity in mice’s brains if oxytocin was the only treatment [82]. This may imply that oxytocin was produced as an auto-immune response to the neuron blockage by toxic beta-amyloid in mice’s brains. Similarly, another animal model study found that oxytocin strengthened social memory [83] and improved spatial memory [84] when mice were in their motherhood. In human studies, it has been reported that oxytocin selectively strengthened participants’ memory for social stimuli depending on the participants’ social contexts and individual attachment styles [84, 85].

A post-mortem study conducted on Alzheimer disease patients’ showed that they had elevated levels of oxytocin in memory-related areas of their brains [86]. However, it was not clear if the elevated levels of oxytocin were caused by more oxytocin receptor expression. Complementary to this study, Takahashi and co-workers found that oxytocin had no influence on synaptic plasticity on its own, but, mixed with beta-amyloid, it did stop the toxic beta-amyloid from negatively affecting synaptic plasticity [82]. Therefore, higher levels of oxytocin in memory-related areas of Alzheimer’s bodies’ brains may be suggesting that, from human auto-immunity perspective, oxytocin was the chemical response to the neuron blockage by the toxic beta-amyloid [86].

Oxytocin has been implicated in so many aspects of social functioning. The therapeutic effects of oxytocin have not only been explored in dementia prevention and treatment, but also have targeted the treatment of diseases for other aberrant social behaviour related disorders [87], such as autism spectrum disorder [88, 89], posttraumatic stress disorder [90], schizophrenia [91], and anxiety disorders [92].

The nature of vascular dementia (VaD) is strongly associated with stroke [93, 94]. To date, there are few therapeutic options to protect cognitive decline arising from cerebrovascular diseases [95, 96]. Worldwide, prevention of strokes and management of post-stroke symptoms have been considered approaches to reduce the vascular dementia initiation [97]. Based on a number of studies in human and animal models, a systematic review conducted by Gutkowska and colleagues concluded that oxytocin has multiple role in protecting cardiovascular system [98], which prevents VaD onset and might be the candidate treatment for VaD [96]. This role includes anti-inflammation, lowering blood pressure, immune-modulation, vasodilatation, antioxidation and wound healing and metabolic moderation [98]. With the reference to the protecting effects of oxytocin in myocardial infarction [99, 100], hepatic ischemia-reperfusion injury [101], renal ischemia/reperfusion injury [102] and cardiomyocyte disorder [103], McKay and co-workers found oxytocin receptor upregulation in postmortem human with VaD through microarray-based profiling and validation [96]. The increased oxytocin receptor expression in peri-infarct regions indicates that oxytocin is the response to microvascular insults. This finding is believed to be a strong evidence of the therapeutic effects of oxytocin on cardiovascular system, and may lead to more research to explore the therapeutic potential of oxytocin on VaD [96].

Frontotemporal dementia (FTD) is an umbrella term for a group of uncommon neurological diseases due to progressive damage to the frontal and/or temporal lobes of the brain. Empathy loss is one of hallmark symptoms of FTD [104, 105]. Oxytocin is an important mediator of social behaviour, potentially enhancing empathy and prosocial behaviours [106]. Finger and colleagues reported that intranasal oxytocin improved the behavioural symptoms in FTD, including levels of apathy and expressions of empathy [106]. And the benefit or efficacy was associated with dosage and time related [106, 107]. This beneficial effect of oxytocin also improved emotional expression processing [108, 109], empathy [110], anxiety [111] and cooperative behaviour [112] in healthy adults and autism patients. Therefore, the mechanism of upregulated oxytocin mediation of empathy and behavioural deficits have been postulated as a potential treatment approach in FTD [106].

### 4.2 Large household creates positive psychological well-being, in particular life purpose to lower dementia risk

Large household promotes more mind-body interaction which offers biological protection against dementia through the therapeutic effects of oxytocin. At the same time, positive psychosocial well-being produced by large household may exert a beneficial slow-down on dementia development [55]. Family members who receive more family support may feel comprehensive positive psychological well-being [55, 113-115]. However, a greater meaning (purpose) of life may be the most important psychological resource to lower dementia risk [55, 113]. Stutin and colleagues explored the protective effects of psychological functioning (life satisfaction, optimism, mastery, purpose in life, positive affect) on preventing high risky population from developing dementia [55]. The results revealed that people from large household showed more psychological well-being leading to lower risk of dementia onset [55]. Interestingly, purpose in life explained 30% dementia risk and this protecting role was independent of the competing effects of other multiple risk factors, such as chronic disease and low physical activities, genetic risk, psychological distress and socioeconomic status [55]. Similarly, five (5) studies conducted by Lambert and co-workers also identified the independent relationship between meaning of life and family support among your people [116].

### 4.3 Social support from household residents promotes healthy lifestyle

Household residents may interact with each other more often to create life satisfaction [26, 117]. They may also share healthcare knowledge, encourage each other to establish healthy lifestyle and utilize health care services in an effective way [118-120]. People with positive psychological well-being tend to practice healthy lifestyle, have more knowledge of health risk factors and attend regular physical examination [121]. The protecting role of such positive psychological well-being has been postulated to decrease the risk in the development of breast cancer and general cancers [121-124].

Large household is even more important for pre-dementia patient, especially the young onset dementia. Generally, young onset dementia present non-specific signs and symptoms at the early onset in young patient, but they are progressing and irreversible [125]. With household residents’ observations and encouragement, the atypical dementia symptoms can be noticed by co-residential members, and accordingly they can have the examination in time, and proper treatment subsequently.

Additionally, from the perspective of evolution, a population with large household offers more chance to survive the natural selection. This population offers the opportunity to have portion of population with less fitness removed without disturbing its activities, for instance, dementia, through greater mortality rate [126-132]. In other words, the genetic background of dementia in such population may be more often eliminated from the population with large household without affecting population as a whole. Therefore, population with large household may have less genes/mutations of dementia to incur high mortality rate of dementia. Furthermore, a population with large household means less birth control and high total fertility rate which allows more biological variation in fertility [133]. A portion of this additional variation, however small, provides the opportunity for the natural selection [133].

Interestingly, large household has shown its consistent and significant, but inverse correlations to dementia mortality rates in the developed world. Several unique phenomena in the developed world can explain this interesting relationship: 1) The fertility rate keeps falling down leading to small household/family size. 2) The cultural doctrines, especially individualism and independence, have reduced the interactions between people living in the developed world. 3) Importantly, dementia care delivery in the developed world is primarily through individual home support service because dementia patients live with small family/household or through nursing homes [134]. While in the developing world, people live with big family/household for most of their life and they would receive the “informal” healthcare from family members and/or household residents [135], instead of the “formal” care through nursing homes.

### 4.5 Strengths and limitations in this study are worth considering

Little work has been done on the dementia epidemiology studies, which may be due to extremely low onset rate for data collection. For example, worldwide, mortality rate of dementia is only 26.61 per 100,000, and this presence is not noticeable. Therefore, the low presence rate of dementia would require unaffordable sample size for identifying small household as the potential risk factor for dementia in individual based epidemiological or laboratory approaches. Ecological studies are based on aggregated quantitative data zooming in the rare presence of dementia 100,000 times, which make dementia presence noticeable for analysing the potential effects of dementia risk-modifying factors at population level. This also suggests the necessity of engaging ecological study into the epidemiology studies of rare chronic disease such as dementia and cancer [42, 43, 126] and Type 1 diabetes [130].

Due to the nature of the cross-sectional data, a couple of intrinsic limitations should be mentioned. Firstly, the results in this study only showed the relationship between household size and dementia mortality rate was correlational, instead of causal. Secondly, the results based on the ecological approach in this work are subject to ecological fallacy. Therefore, the protective role of large household size may not always hold true for each individual to predict their dementia specific death risk. A further limitation of this work is that the data employed in this study might be crude. The WHO, United Nations and World Bank may have made some random errors arising from the methodologies used for collecting and aggregating the data.

Regardless of the strength and limitations of the data quality, we have showed that countries with large household size had lower dementia mortality rate in different data analysis models. The findings in this study may have shed light for further research into the subject with exposure based longitudinal cohort studies at population level. Accordingly, this will lead to far reaching public health implications in dementia and its prevention.

## 5. Conclusions

Independent of ageing, urban lifestyle and low socio-economic status, large household has been shown its significant protective role against dementia mortality in this study. This forward-thinking study approach will help shed light on the further study into the protective role of positive psychological well-being against dementia. As part of dementia prevention, healthcare practitioners should encourage people to increase their positive interactions with people from their neighbourhood or other fields when large household/family size is impossible to achieve.

## Data Availability

The data sources have been described in the section of Materials and Methods. All data for this study are freely available from the United Nation agencies official websites. The formal permission to use the data for non-commercial purpose is not necessary as it is compliant with the agencys public permission in their terms and conditions.

## List of abbreviations

GDP: Gross Domestic Product
Life e(60): Life expectancy at 60 years old
SES: Socioeconomic status
UN: United Nations
WHO: World Health Organization
DMR: Dementia mortality rate

## Ethics approval and informed consent

All the data supporting our findings in this paper were freely downloaded from the UN agencies’ websites. No ethical approval or written informed consent for participation was required.

## Funding

There is no specific funding related to this study.

## Authors’ contributions

WY Conceptualization; Data curation; Formal analysis; Investigation; Methodology;

Roles/Writing - original draft; Writing - review & editing.

MH before interpreting the analysis results. WY drafted the text with contributions from MH Conceptualization; Formal analysis; Investigation; Methodology; Validation; Visualization; Writing - review & editing.

## Acknowledgments

The authors express appreciation to Cherian Varghese from the Management of on communicable

Diseases, Disability, Violence and Injury Prevention Department (NVI) of World Health Organization for the assistance in locating the data.

## Declarations

### Consent for publication

Not applicable.

### Availability of data and materials

The data sources have been described in the section of “Materials and Methods”. All data for this study are freely available from the United Nation agencies’ official websites. The formal permission to use the data for non-commercial purpose is not necessary as it is compliant with the agency’s public permission in their terms and conditions.

### Competing interest

The authors declared that there is no conflict of interest.

## Notes

### Competing Interest Statement

The authors have declared no competing interest.

### Funding Statement

This study did not receive any funding.

## References

1. Burns A: iliffe S. Dementia Bmj 2009, 338:b75.

2. World Health Organization: Dementia fact sheet In: https://www.whoint/news-room/fact-sheets/detail/dementia. WHO Official Website: WHO; September 21, 2020.

3. Burns A, Iliffe S: Dementia. BMJ : British Medical Journal (Online) 2009, 338(7691).

4. World Health Organization: The Epididemiology and Impact of Dementia. In. Online; 2015.

5. Prince M, Comas-Herrera A, Knapp M, Guerchet M, Karagiannidou M: World Alzheimer report 2016: improving healthcare for people living with dementia: coverage, quality and costs now and in the future. 2016.

6. Care AsANP, Workgroup SM, Borson S, Boustani MA, Buckwalter KC, Burgio LD, Chodosh J, Fortinsky RH, Gifford DR, Gwyther LP et al: Report on milestones for care and support under the US National Plan to Address Alzheimer’s Disease. Alzheimer’s & Dementia 2016, 12(3):334–369.

7. Kmietowicz Z: Cameron launches challenge to end “national crisis” of poor dementia care. In.: British Medical Journal Publishing Group; 2012.

8. van der Flier WM, Scheltens P: Epidemiology and risk factors of dementia. Journal of Neurology, Neurosurgery & Psychiatry 2005, 76(Suppl 5):v2–v7.

9. Loy CT, Schofield PR, Turner AM, Kwok JB: Genetics of dementia. The Lancet 2014, 383(9919):828–840.

10. Roccisano D, Henneberg M, Saniotis A: A possible cause of Alzheimer’s dementia– Industrial soy foods. Medical hypotheses 2014, 82(3):250–254.

11. Zis P, Hadjivassiliou M: Treatment of neurological manifestations of gluten sensitivity and coeliac disease. Current treatment options in neurology 2019, 21(3):10.

12. Makhlouf S, Messelmani M, Zaouali J, Mrissa R: Cognitive impairment in celiac disease and non-celiac gluten sensitivity: Review of literature on the main cognitive impairments, the imaging and the effect of gluten free diet. Acta Neurologica Belgica 2018, 118(1):21–27.

13. Burckhardt M, Herke M, Wustmann T, Watzke S, Langer G, Fink A: Omega-3 fatty acids for the treatment of dementia. Cochrane Database of Systematic Reviews 2016(4).

14. Saczynski JS, Beiser A, Seshadri S, Auerbach S, Wolf P, Au R: Depressive symptoms and risk of dementia: the Framingham Heart Study. Neurology 2010, 75(1):35–41.

15. Terracciano A, Sutin AR, An Y, O’Brien RJ, Ferrucci L, Zonderman AB, Resnick SM: Personality and risk of Alzheimer’s disease: new data and meta-analysis. Alzheimer’s & Dementia 2014, 10(2):179–186.

16. Badcock CR: Evolutionary psychology: A critical introduction: Macmillan International Higher Education; 2000.

17. Clark G, Henneberg M: The life history of Ardipithecus ramidus: a heterochronic model of sexual and social maturation. Anthropological Review 2015, 78(2).

18. K H, RR P: The Evolution of Human Life History. Santa Fe: School of American Research Press; 2006.

19. Isler K, van Schaik CP: Allomaternal care, life history and brain size evolution in mammals. J Hum Evol 2012, 63(1):52–63.

20. Henneberg M, Saniotis A: The Dynamic Human. Sharjah, U.A.E.: Bentham Science Publishers Ltd.; 2016.

21. Crawford C, Krebs D: Foundations of evolutionary psychology: Psychology Press; 2012.

22. Grinde B: Can the concept of discords help us find the causes of mental diseases? Medical hypotheses 2009, 73(1):106–109.

23. Grinde B: The biology of happiness: Springer Science & Business Media; 2012.

24. Howell RT, Kern ML, Lyubomirsky S: Health benefits: Meta-analytically determining the impact of well-being on objective health outcomes. Health Psychology Review 2007, 1(1):83–136.

25. Hughes ME, Waite LJ: Health in household context: Living arrangements and health in late middle age. Journal of health and social behavior 2002, 43(1):1.

26. Nan H, Ni MY, Lee PH, Tam WW, Lam TH, Leung GM, McDowell I: Psychometric evaluation of the Chinese version of the Subjective Happiness Scale: evidence from the Hong Kong FAMILY Cohort. International journal of behavioral medicine 2014, 21(4):646–652.

27. You W, Rühli FJ, Henneberg RJ, Henneberg M: Greater family size is associated with less cancer risk: an ecological analysis of 178 countries. BMC cancer 2018, 18(1):924.

28. Van Campen C, Iedema J: Are persons with physical disabilities who participate in society healthier and happier? Structural equation modelling of objective participation and subjective well-being. Quality of Life Research 2007, 16(4):635.

29. Mukuria C, Brazier J: Valuing the EQ-5D and the SF-6D health states using subjective well-being: a secondary analysis of patient data. Social Science & Medicine 2013, 77:97–105.

30. Grinde B, Tambs K: Effect of household size on mental problems in children: results from the Norwegian Mother and Child Cohort study. BMC psychology 2016, 4(1):31.

31. World Health Organization: Global Health Estimates 2016: Deaths by Cause, Age, Sex, by Country and by Region, 2000-2016. In. Geneva; 2018.

32. World Health Organization: WHO methods and data sources for global causes of death 2000-2016. Global Health Estimates Technical Paper WHO/HIS/IER/GHE/2018.3.. In. Geneva; 2018

33. United Nations DoEaSA, Population Division; : Household Size and Composition Around the World 2017 – Data Booklet (ST/ESA/SER.A/405); 2017.

34. Casper LM, Bryson K: Current Population Reports: Household and Family Characteristics: March 1998 (Update)(Publication no. P20–515). Washington, DC: US Bureau of the Census 1998.

35. Lugaila TA: Marital Status and Living Arrangements, March 1998 (Update): US Bureau of the Census; 1998.

36. Australian Bureau of Statistics: Household and Family Projections, Australia. In.; 14/03/2019.

37. Organization;; TWH: Healthy life expectancy (HALE). In.; 2010.

38. Speechly CM, Bridges-Webb C, Passmore E: The pathway to dementia diagnosis. Medical Journal of Australia 2008, 189(9):487–489.

39. Rubinow IM: Social insurance: With special reference to American conditions: Henry Holt & Co., New York, 1913.; 1913.

40. World Bank Open Data [http://data.worldbank.org/]

41. WHO: Diet, nutrition and the prevention of chronic diseases: report of a joint WHO/FAO expert consultation (WHO technical report series ; 916). In. Geneva; 2003.

42. You W, Symonds I, Henneberg M: Low fertility may be a significant determinant of ovarian cancer worldwide: an ecological analysis of cross-sectional data from 182 countries. Journal of ovarian research 2018, 11(1):68.

43. You W, Symonds I, Rühli FJ, Henneberg M: Decreasing birth rate determining worldwide incidence and regional variation of female breast Cancer. Advances in Breast Cancer Research 2018, 7(01):1–14.

44. Country and Lending Groups | Data [http://data.worldbank.org/about/country-and-lending-groups]

45. WHO regional offices [http://www.who.int]

46. Member Countries [http://www.acddialogue.com]

47. Member Economies-Asia-Pacific Economic Cooperation [http://www.apec.org]

48. UNESCO Regions-Latin America and the Caribbean [http://www.unesco.org]

49. Southern African Development Community: Member States [http://www.sadc.int]

50. Cadar D: A Life Course Approach to Dementia Prevention. Journal of Aging and Geriatric Medicine 2017, 1(2).

51. Ritchie CW, Terrera GM, Quinn TJ: Dementia trials and dementia tribulations: methodological and analytical challenges in dementia research. Alzheimer’s research & therapy 2015, 7(1):31.

52. Samsi K, Manthorpe J: Care pathways for dementia: current perspectives. Clinical Interventions in Aging 2014, 9:2055.

53. Krause N, Bengston V, Silverstein M, Putney N, Gans D: Handbook of Theories of Aging; 2009.

54. Zunzunegui M-V, Béland F, Sanchez M-T, Otero A: Longevity and relationships with children: the importance of the parental role. BMC public health 2009, 9(1):351.

55. Sutin AR, Stephan Y, Terracciano A: Psychological well-being and risk of dementia. International journal of geriatric psychiatry 2018, 33(5):743–747.

56. Boyle PA, Buchman AS, Barnes LL, Bennett DA: Effect of a purpose in life on risk of incident Alzheimer disease and mild cognitive impairment in community-dwelling older persons. Archives of general psychiatry 2010, 67(3):304–310.

57. Wang H-X, Karp A, Winblad B, Fratiglioni L: Late-life engagement in social and leisure activities is associated with a decreased risk of dementia: a longitudinal study from the Kungsholmen project. American journal of epidemiology 2002, 155(12):1081–1087.

58. Marioni RE, Proust-Lima C, Amieva H, Brayne C, Matthews FE, Dartigues J-F, Jacqmin-Gadda H: Social activity, cognitive decline and dementia risk: a 20-year prospective cohort study. BMC public health 2015, 15(1):1–8.

59. Bechara A, Naqvi N: Listening to your heart: interoceptive awareness as a gateway to feeling. Nature neuroscience 2004, 7(2):102–103.

60. Cacioppo JT, Berntson GG, Sheridan JF, McClintock MK: Multilevel integrative analyses of human behavior: Social neuroscience and the complementing nature of social and biological approaches. Psychological bulletin 2000, 126(6):829.

61. Damasio A, Carvalho GB: The nature of feelings: evolutionary and neurobiological origins. Nature reviews neuroscience 2013, 14(2):143–152.

62. Lee Y-S, Ryu Y, Jung W-M, Kim J, Lee T, Chae Y: Understanding mind-body interaction from the perspective of east Asian medicine. Evidence-Based Complementary and Alternative Medicine 2017, 2017.

63. James W: II.—WHAT IS AN EMOTION? Mind 1884, os-IX(34).

64. Huangdi, Qibo, Other ministers: Huangdi Neijing. Wuhan: Wuhan Publishing House; 2013.

65. Magon N, Kalra S: The orgasmic history of oxytocin: Love, lust, and labor. Indian journal of endocrinology and metabolism 2011, 15(Suppl3):S156.

66. Takayanagi Y, Yoshida M, Bielsky IF, Ross HE, Kawamata M, Onaka T, Yanagisawa T, Kimura T, Matzuk MM, Young LJ: Pervasive social deficits, but normal parturition, in oxytocin receptor-deficient mice. Proceedings of the National Academy of Sciences of the United States of America 2005, 102(44):16096–16101.

67. White-Traut R, Watanabe K, Pournajafi-Nazarloo H, Schwertz D, Bell A, Carter CS: Detection of salivary oxytocin levels in lactating women. Developmental psychobiology 2009, 51(4):367–373.

68. Carmichael MS, Humbert R, Dixen J, Palmisano G, Greenleaf W, Davidson JM: Plasma oxytocin increases in the human sexual response. The Journal of Clinical Endocrinology & Metabolism 1987, 64(1):27–31.

69. Carmichael MS, Warburton VL, Dixen J, Davidson JM: Relationships among cardiovascular, muscular, and oxytocin responses during human sexual activity. Archives of sexual behavior 1994, 23(1):59–79.

70. Gordon Jr G, Burch RL, Platek SM: Does semen have antidepressant properties? Archives of Sexual Behavior 2002, 31(3):289–293.

71. Kendrick KM: The neurobiology of social bonds. Journal of neuroendocrinology 2004, 16(12):1007–1008.

72. Weisman O, Zagoory-Sharon O, Feldman R: Oxytocin administration to parent enhances infant physiological and behavioral readiness for social engagement. Biological psychiatry 2012, 72(12):982–989.

73. MacDonald K, MacDonald TM: The peptide that binds: a systematic review of oxytocin and its prosocial effects in humans. Harvard review of psychiatry 2010, 18(1):1–21.

74. Van IJzendoorn MH, Bakermans-Kranenburg MJ: A sniff of trust: meta-analysis of the effects of intranasal oxytocin administration on face recognition, trust to in-group, and trust to out-group. Psychoneuroendocrinology 2012, 37(3):438–443.

75. Wudarczyk OA, Earp BD, Guastella A, Savulescu J: Could intranasal oxytocin be used to enhance relationships? Research imperatives, clinical policy, and ethical considerations. Current opinion in psychiatry 2013, 26(5):474.

76. Insel TR, Hulihan TJ: A gender-specific mechanism for pair bonding: oxytocin and partner preference formation in monogamous voles. Behavioral neuroscience 1995, 109(4):782.

77. Young LJ, Murphy Young AZ, Hammock EA: Anatomy and neurochemistry of the pair bond. Journal of Comparative Neurology 2005, 493(1):51–57.

78. Angeles L: Children and life satisfaction. Journal of happiness Studies 2010, 11(4):523–538.

79. Churchland PS, Winkielman P: Modulating social behavior with oxytocin: how does it work? What does it mean? Hormones and behavior 2012, 61(3):392–399.

80. Olff M, Frijling JL, Kubzansky LD, Bradley B, Ellenbogen MA, Cardoso C, Bartz JA, Yee JR, Van Zuiden M: The role of oxytocin in social bonding, stress regulation and mental health: an update on the moderating effects of context and interindividual differences. Psychoneuroendocrinology 2013, 38(9):1883–1894.

81. Balmus I, Ciobica A, Stoica B, Lefter R, Cojocaru S, Reznikov A: Effects of oxytocin administration on oxidative markers in the temporal lobe of aged rats. Neurophysiology 2019, 51(1):18–24.

82. Takahashi J, Yamada D, Ueta Y, Iwai T, Koga E, Tanabe M, Oka J-I, Saitoh A: Oxytocin reverses Aβ-induced impairment of hippocampal synaptic plasticity in mice. Biochemical and biophysical research communications 2020.

83. Guzmán YF, Tronson NC, Jovasevic V, Sato K, Guedea AL, Mizukami H, Nishimori K, Radulovic J: Fear-enhancing effects of septal oxytocin receptors. Nature neuroscience 2013, 16(9):1185–1187.

84. Tomizawa K, Iga N, Lu Y-F, Moriwaki A, Matsushita M, Li S-T, Miyamoto O, Itano T, Matsui H: Oxytocin improves long-lasting spatial memory during motherhood through MAP kinase cascade. Nature neuroscience 2003, 6(4):384–390.

85. Heinrichs M, Meinlschmidt G, Wippich W, Ehlert U, Hellhammer DH: Selective amnesic effects of oxytocin on human memory. Physiology & behavior 2004, 83(1):31–38.

86. Mazurek MF, Beal MF, Bird ED, Martin JB: Oxytocin in Alzheimer’s disease: postmortem brain levels. Neurology 1987, 37(6):1001–1001.

87. Fineberg SK, Ross DA: Oxytocin and the social brain. Biological psychiatry 2017, 81(3):e19.

88. Parker KJ, Oztan O, Libove RA, Sumiyoshi RD, Jackson LP, Karhson DS, Summers JE, Hinman KE, Motonaga KS, Phillips JM: Intranasal oxytocin treatment for social deficits and biomarkers of response in children with autism. Proceedings of the National Academy of Sciences 2017, 114(30):8119–8124.

89. Alvares GA, Quintana DS, Whitehouse AJ: Beyond the hype and hope: critical considerations for intranasal oxytocin research in autism spectrum disorder. Autism Research 2017, 10(1):25–41.

90. Knobloch HS, Charlet A, Hoffmann LC, Eliava M, Khrulev S, Cetin AH, Osten P, Schwarz MK, Seeburg PH, Stoop R: Evoked axonal oxytocin release in the central amygdala attenuates fear response. Neuron 2012, 73(3):553–566.

91. Oya K, Matsuda Y, Matsunaga S, Kishi T, Iwata N: Efficacy and safety of oxytocin augmentation therapy for schizophrenia: an updated systematic review and meta-analysis of randomized, placebo-controlled trials. European archives of psychiatry and clinical neuroscience 2016, 266(5):439–450.

92. Dodhia S, Hosanagar A, Fitzgerald DA, Labuschagne I, Wood AG, Nathan PJ, Phan KL: Modulation of resting-state amygdala-frontal functional connectivity by oxytocin in generalized social anxiety disorder. Neuropsychopharmacology 2014, 39(9):2061–2069.

93. Mijajlović MD, Pavlović A, Brainin M, Heiss W-D, Quinn TJ, Ihle-Hansen HB, Hermann DM, Assayag EB, Richard E, Thiel A: Post-stroke dementia–a comprehensive review. BMC medicine 2017, 15(1):1–12.

94. Chen A, Akinyemi RO, Hase Y, Firbank MJ, Ndung’u MN, Foster V, Craggs LJ, Washida K, Okamoto Y, Thomas AJ: Frontal white matter hyperintensities, clasmatodendrosis and gliovascular abnormalities in ageing and post-stroke dementia. Brain 2016, 139(1):242–258.

95. Attems J, Jellinger KA: The overlap between vascular disease and Alzheimer’s disease-lessons from pathology. BMC medicine 2014, 12(1):206.

96. McKay EC, Beck JS, Khoo SK, Dykema KJ, Cottingham SL, Winn ME, Paulson HL, Lieberman AP, Counts SE: Peri-Infarct Upregulation of the Oxytocin Receptor in Vascular Dementia. Journal of Neuropathology & Experimental Neurology 2019, 78(5):436–452.

97. Baskys A, Cheng J-x: Pharmacological prevention and treatment of vascular dementia: approaches and perspectives. Experimental gerontology 2012, 47(11):887–891.

98. Gutkowska J, Jankowski M: Oxytocin revisited: its role in cardiovascular regulation. Journal of neuroendocrinology 2012, 24(4):599–608.

99. Jankowski M, Bissonauth V, Gao L, Gangal M, Wang D, Danalache B, Wang Y, Stoyanova E, Cloutier G, Blaise G: Anti-inflammatory effect of oxytocin in rat myocardial infarction. Basic Research in Cardiology 2010, 105(2):205–218.

100. Petersson M, Lundeberg T, Sohlström A, Wiberg U, Uvnäs-Moberg K: Oxytocin increases the survival of musculocutaneous flaps. Naunyn-Schmiedeberg’s archives of pharmacology 1998, 357(6):701–704.

101. Düşünceli F, Işeri SÖ, Ercan F, Gedik N, Yeğen C, Yeğen BÇ: Oxytocin alleviates hepatic ischemia–reperfusion injury in rats. Peptides 2008, 29(7):1216–1222.

102. Tuğtepe H, şener G, Biyikli NK, Yüksel M, Çetinel ş, Gedik N, Yeğen BÇ: The protective effect of oxytocin on renal ischemia/reperfusion injury in rats. Regulatory peptides 2007, 140(3):101–108.

103. Gonzalez-Reyes A, Menaouar A, Yip D, Danalache B, Plante E, Noiseux N, Gutkowska J, Jankowski M: Molecular mechanisms underlying oxytocin-induced cardiomyocyte protection from simulated ischemia–reperfusion. Molecular and cellular endocrinology 2015, 412:170–181.

104. Neary D, Snowden JS, Gustafson L, Passant U, Stuss D, Black S, Freedman M, Kertesz A, Robert P, Albert M: Frontotemporal lobar degeneration: a consensus on clinical diagnostic criteria. Neurology 1998, 51(6):1546–1554.

105. Rankin KP, Kramer JH, Miller BL: Patterns of cognitive and emotional empathy in frontotemporal lobar degeneration. Cognitive and Behavioral Neurology 2005, 18(1):28–36.

106. Finger EC, MacKinley J, Blair M, Oliver LD, Jesso S, Tartaglia MC, Borrie M, Wells J, Dziobek I, Pasternak S: Oxytocin for frontotemporal dementia: a randomized dose-finding study of safety and tolerability. Neurology 2015, 84(2):174–181.

107. Jesso S, Morlog D, Ross S, Pell MD, Pasternak SH, Mitchell DG, Kertesz A, Finger EC: The effects of oxytocin on social cognition and behaviour in frontotemporal dementia. Brain 2011, 134(9):2493–2501.

108. Hollander E, Bartz J, Chaplin W, Phillips A, Sumner J, Soorya L, Anagnostou E, Wasserman S: Oxytocin increases retention of social cognition in autism. Biological psychiatry 2007, 61(4):498–503.

109. Marsh AA, Henry HY, Pine DS, Blair R: Oxytocin improves specific recognition of positive facial expressions. Psychopharmacology 2010, 209(3):225–232.

110. Hurlemann R, Patin A, Onur OA, Cohen MX, Baumgartner T, Metzler S, Dziobek I, Gallinat J, Wagner M, Maier W: Oxytocin enhances amygdala-dependent, socially reinforced learning and emotional empathy in humans. Journal of Neuroscience 2010, 30(14):4999–5007.

111. Gamer M, Zurowski B, Büchel C: Different amygdala subregions mediate valence-related and attentional effects of oxytocin in humans. Proceedings of the National Academy of Sciences 2010, 107(20):9400–9405.

112. Kosfeld M, Heinrichs M, Zak PJ, Fischbacher U, Fehr E: Oxytocin increases trust in humans. Nature 2005, 435(7042):673–676.

113. Symister P, Friend R: The influence of social support and problematic support on optimism and depression in chronic illness: A prospective study evaluating self-esteem as a mediator. Health Psychology 2003, 22(2):123.

114. Thoits PA: Stress and health: Major findings and policy implications. Journal of health and social behavior 2010, 51(1_suppl):S41–S53.

115. Fukukawa Y, Tsuboi S, Niino N, Ando F, Kosugi S, Shimokata H: Effects of social support and self-esteem on depressive symptoms in Japanese middle-aged and elderly people. Journal of epidemiology 2000, 10(1sup):63–69.

116. Lambert KM, Barry P, Stokes G: Risk management and legal issues with the use of social media in the healthcare setting. Journal of Healthcare Risk Management 2012, 31(4):41–47.

117. Angeles L: Children and Life Satisfaction. Journal of Happiness Studies 2009, 11(4):523–538.

118. Cohen S: Social relationships and health. American psychologist 2004, 59(8):676.

119. Reczek C, Beth Thomeer M, Lodge AC, Umberson D, Underhill M: Diet and exercise in parenthood: A social control perspective. Journal of Marriage and Family 2014, 76(5):1047–1062.

120. Aizer AA, Chen MH, McCarthy EP, Mendu ML, Koo S, Wilhite TJ, Graham PL, Choueiri TK, Hoffman KE, Martin NE et al: Marital status and survival in patients with cancer. Journal of clinical oncology : official journal of the American Society of Clinical Oncology 2013, 31(31):3869–3876.

121. Bai A, Li H, Huang Y, Liu X, Gao Y, Wang P, Dai H, Song F, Hao X, Chen K: A survey of overall life satisfaction and its association with breast diseases in Chinese women. Cancer medicine 2016, 5(1):111–119.

122. Kim Y, Duhamel KN, Valdimarsdottir HB, Bovbjerg DH: Psychological distress among healthy women with family histories of breast cancer: effects of recent life events. Psycho-oncology 2005, 14(7):555–563.

123. Peled R, Carmil D, Siboni-Samocha O, Shoham-Vardi I: Breast cancer, psychological distress and life events among young women. BMC cancer 2008, 8:245.

124. You W, Henneberg M: Cancer incidence increasing globally: The role of relaxed natural selection. Evol Appl 2017, 00:1–13.

125. Elhusein B, Radwan A, Khairi A, Ahmed M: Atypical presentation of progressive young onset dementia. BMJ Case Reports CP 2018, 11(1).

126. You W, M H: Cancer incidence increasing globally: The role of relaxed natural selection. Evol Appl 2017, 00:1–13.

127. Budnik A, Henneberg M: Worldwide Increase of Obesity is Related to the Reduced Opportunity for Natural Selection. PloS one 2017, 12(1).

128. Henneberg M, Piontek J: Biological state index of human groups. Przeglad Anthropologiczny 1975, XLI:191–201.

129. Henneberg M: Reproductive possibilities and estimations of the biological dynamics of earlier human populations. Journal of Human Evolution 1976, 5:41–48.

130. You W-P, Henneberg M: Type 1 diabetes prevalence increasing globally and regionally: the role of natural selection and life expectancy at birth. BMJ Open Diabetes Research & Care 2016, 4(1):e000161.

131. Rühli F, Henneberg M: Biological future of humankind – ongoing evolution and the impact of recognition of human biological variation. In: On Human Nature Biology, Psychology, Ethics, Politics, and Religion 2016. edn. Edited by Tibayrenc M, Ayala FJ. eBook: Academic Press: Elsevier; 2016: 263–275.

132. Stephan CN, Henneberg M: Medicine may be reducing the human capacity to survive. Medical hypotheses 2001, 57(5):633–637.

133. Henneberg M: Natural selection through differential fertility in human populations: Quantitative evaluation of selection intensity. Przeglad Antropologiczny 1980, 46:21–60.

134. Brodaty H, Donkin M: Family caregivers of people with dementia. Dialogues in clinical neuroscience 2009, 11(2):217.

135. Prince M: Care arrangements for people with dementia in developing countries. International journal of geriatric psychiatry 2004.

